# An Imputation-Based Approach for Augmenting Sparse Epidemiological Signals

**DOI:** 10.1101/2024.07.31.24311314

**Authors:** Amy E. Benefield, Desiree Williams, VP Nagraj

## Abstract

Near-term disease forecasting and scenario projection efforts rely on the availability of data to train and evaluate model performance. In most cases, more extensive epidemiological time series data can lead to better modeling results and improved public health insights. Here we describe a procedure to augment an epidemiological time series. We used reported flu hospitalization data from FluSurv-NET, the National Healthcare Safety Network, and flu outpatient visits from ILINet to estimate a complete time series of flu hospitalization counts dating back to 2009. The augmentation process includes concatenation, interpolation, extrapolation, and imputation steps, each designed to address specific data gaps. We demonstrate the forecasting performance gain when the extended time series is used to train flu hospitalization models at the state and national level.

## 1 Background

Operational disease modeling initiatives can provide key information for public health response. However, limited availability of historical epidemiological data pose a challenge for training accurate disease forecasting and projection models. As contributors to the CDC FluSight consortium, we have experienced these challenges as they pertain to forecasting near-term influenza hospitalizations in the United States. In 2020, the U.S. Department of Health and Human Services (HHS) began reporting flu hospitalizations through the HHS Protect network, which is now known as the National Healthcare Safety Network (NHSN). Because the field was only mandatory between February 2022 and April 2024 (U.S. Department of Health and Human Services 2023), there are just over two years worth of the gold standard, state-level flu hospitalization surveillance data. More training data can improve the performance of time series models and enable the use of new methods. We therefore sought to augment the NHSN data to estimate flu hospitalizations prior to 2020. What follows is a description of the approach we developed to extend this time series using additional data signals to create a continuous, comprehensive estimate of flu hospitalizations in the United States.

## 2 Methods

### 2.1 Data Sources

To extend the flu hospitalization time series, we considered two orthogonal flu reporting mechanisms: FluSurv-NET (FSN) and ILINet. While flu surveillance systems such as FSN and ILINet have critical public health utility, they are known to, in some cases, exhibit population bias, limited geographic representation, and suboptimal provider coverage (Scarpino et al. 2020; Scarpino, Dimitrov, and Meyers 2012; Chaves et al. 2015). FSN provides flu hospitalization records from selected states, with data from some states dating back as far as 2009 (Centers for Disease Control and Prevention 2023). ILINet is a national, state-level record of outpatient influenza-like illness (ILI) visits. It does not track hospitalizations, and the dataset may include patient visits caused by other respiratory diseases that produce similar symptoms (Centers for Disease Control and Prevention 2024).

### 2.2 Data Augmentation Process

The data augmentation process that we developed consists of four steps (concatenation, interpolation, extrapolation, and imputation), with the resulting signal intended to approximate NHSN flu hospitalization counts. While we used available NHSN data where possible, we aimed to minimize bias by leveraging alternative signals for flu hospitalizations (FSN) and ILI (ILINet) at the appropriate steps. We began by concatenating data from the NHSN and FSN signals, both of which report state-level hospitalizations. We then interpolated hospitalization counts between the end of the previous season and start of the following season to account for gaps in FSN reporting. With the NHSN and FSN data aligned from the concatenation and interpolation steps, we trained a model to extrapolate hospitalization counts from the FSN hospitalization rates. As a last step, we combined the available NHSN reporting, extrapolated hospitalization counts, and ILI data as inputs to a multiple imputation algorithm.

#### 2.2.1 Concatenation

We began by retrieving and reconciling available flu hospitalization signals. Using the Epidata API R package (epidatr) (Farrow et al. 2015), we obtained FSN reporting values from the 19 participating states, some with records dating back to 2009. The package has a cutoff of April 25th, 2020, so we manually downloaded FSN data between October 3rd, 2020 and April 21st, 2024. FSN does not report during non-seasonal weeks, creating a gap between April and October. FSN has two entries for New York State (NY Albany and NY Rochester), and we averaged these values to summarize New York hospitalizations. For downstream analyses, we used the usa R package (Nicholls 2024) to annotate the data with 2019 state populations, converting rates per 100k to counts (the unit used by NHSN). Further, we mapped NHSN regions and the proportion of the state represented by FSN to the individual states (“Influenza Hospitalization Surveillance Network (FluSurv-NET)” 2023; U.S. Department of Health and Human Services 2024). Using the fiphde R package (Nagraj et al. 2024), we queried all state-level hospitalization data from NHSN, removed DC, and merged all datasets. Finally, we added rows for all missing dates for each state dating back to 2009. The combined FSN and NHSN dataset from the concatenation step is depicted in Figure 1. However, at this point the data has many missing entries for flu hospitalizations, both from states that never reported to FSN (e.g., Alaska in Figure 1) and from states that did but had gaps in reporting during the summer months and during the pandemic (e.g., California in Figure 1). Note that the concatenation described here was implemented to reconcile only hospitalization signals. ILI data from ILINet were used as a covariate in the imputation algorithm, and were retrieved and cleaned prior to the imputation step.

**Figure 1:**
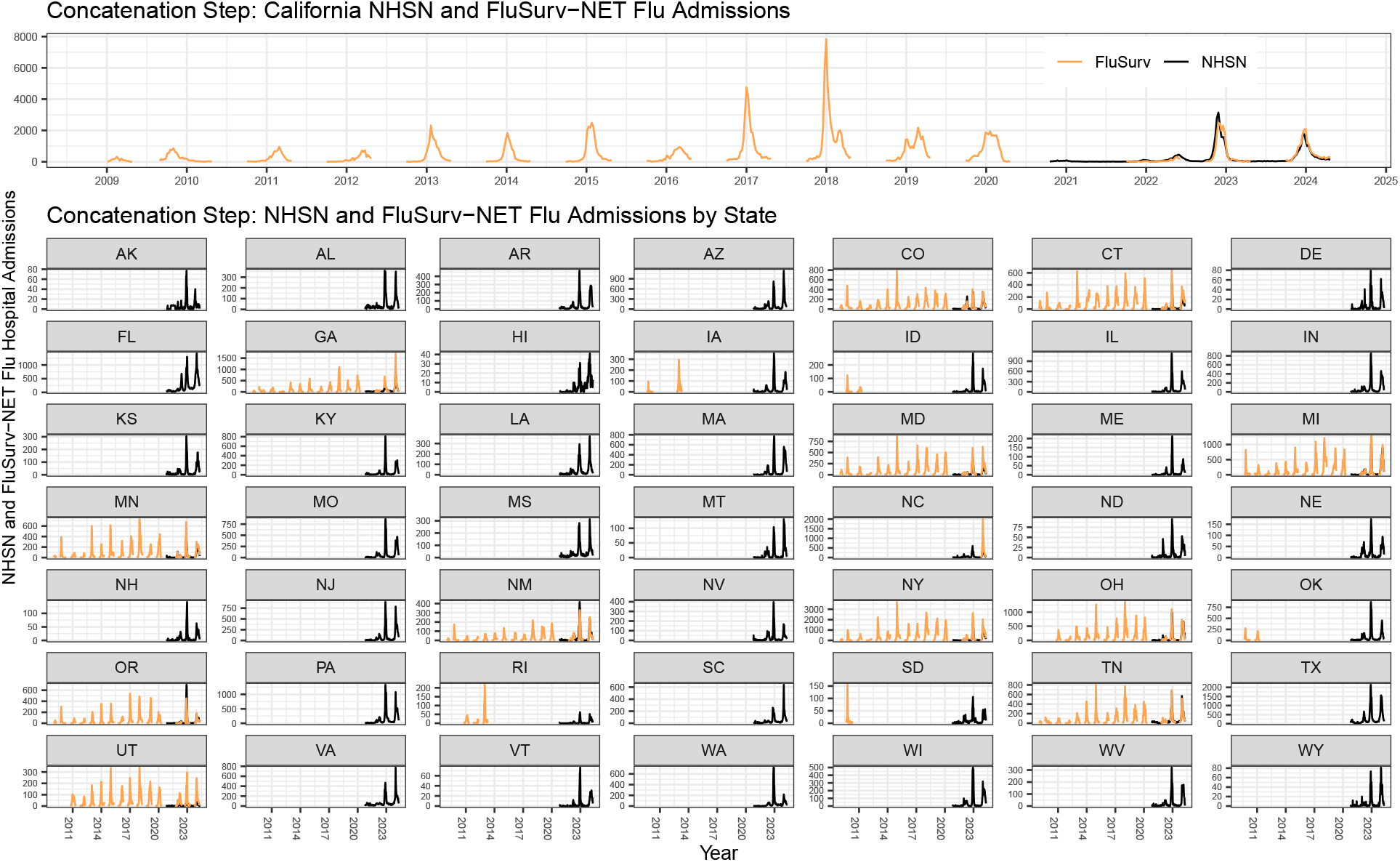
Concatenation of FluSurv-NET and NHSN hospitalization records. Many states have never participated in FSN (orange), and thus only the NHSN data (black) are shown. Some states have reported to FSN intermittently (e.g., Iowa) or do not have records dating back to 2009 (e.g., Utah). California (top panel) was used as an exemplar state, as it has a long history of reporting to FSN. Still, there are no records for non-seasonal weeks (e.g., gaps in the FSN record for California).

#### 2.2.2 Interpolation

The first missing data that we addressed were the short gaps in FSN hospitalization records across non-seasonal weeks and some occasional missed reports. Because these gaps were either very short (e.g., intermittent missed weeks) or across summer months when flu hospitalizations tend to be low, we employed a simple linear interpolation at the state level. Some states that have historically reported to FSN had long periods spanning several years with no reporting. To avoid estimating large spans of missing data with linear interpolation, we limited interpolation to gaps of less than 26 weeks. Figure 2 demonstrates the interpolated values alongside the available FSN observations.

**Figure 2:**
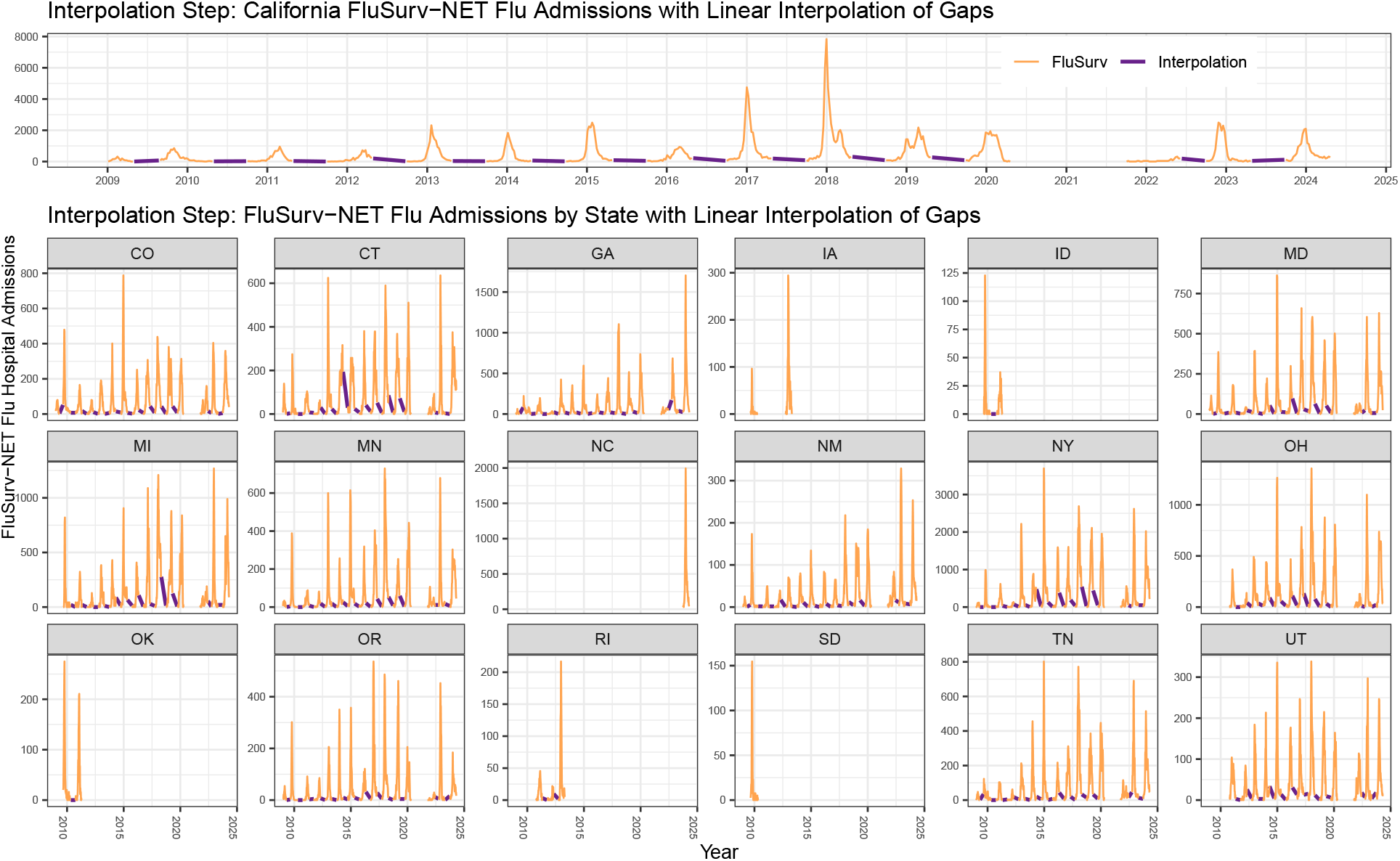
FluSurv-NET hospitalization records with linear interpolation. All states reporting to FSN had gaps during non-seasonal weeks, and we used linear interpolation (purple lines) to estimate hospitalizations during those short gaps. Note that we did not interpolate across gaps longer than 26 weeks, such as the multi-year gaps in Iowa. Further, no states reported to FSN during the pandemic, which is clearly shown in California (top panel) from early 2020 to late 2021.

#### 2.2.3 Extrapolation

After the interpolation step, the dataset still contains many gaps but now includes two fields for hospitalizations for select states: one from FSN and the other from NHSN. We planned to use a multivariate imputation procedure downstream to complete the dataset for all states and weeks. Imputation procedures can be heavily biased by including collinear predictors (Van Buuren and Groothuis-Oudshoorn 2011), and we therefore sought to consolidate the two variables representing hospitalizations. Though the FSN record and the ground-truth NHSN data are highly correlated when both data are present, they are not identical signals. When translated to counts, FSN hospitalizations tend to be higher than the values reported by NHSN for the same state and date. As such, we established a conversion equation to extrapolate the interpolated FSN data to NHSN data. Because the ILINet data records outpatient ILI visits and not hospitalizations, we did not need to perform a similar consolidation procedure.

Our extrapolation was based on a generalized linear model (GLM) trained on instances when both NHSN and FSN data were present. We used this model to predict NHSN hospitalizations when only FSN data were present, resulting in a single variable for hospital admissions. Fourteen of the nineteen FSN-participating states had overlapping NHSN records, with a total of 1,388 data points, of which, we excluded five outliers that were identified from diagnostic plots of the GLM. Using Akaike information criterion (AIC), Bayesian information criterion (BIC) and log likelihood, we determined that the best model formulation was a Gaussian GLM with NHSN flu admissions as the response variable, FSN hospitalizations (*β* = 0.818, SE = 0.008, p < 0.001) as the primary predictor variable, and population size (*β* = 6.732e-07, SE = 1.543e-07, p < 0.001) and NHSN-defined region (*β* = 0.864, SE = 0.567, p = 0.128) as covariates (see Supplementary Figure S1 for a fit of the model). Though they were significant terms when included in the model, we excluded location (i.e., state), the proportion of the population represented by FSN, and the NHSN flu admission coverage rate as covariates, because these variables were present in the data used to train the GLM but not always in the FSN data used for extrapolation. Date, epiyear, and epiweek were not significant terms and were excluded; the relationship between NHSN and FSN hospital admissions did not vary with time. We extrapolated a total of 7,862 FSN hospital admissions, and in cases where both data were present, we kept the NHSN record rather than the extrapolated value. Figure 3 shows the reported, interpolated, and extrapolated values for the states reporting to FSN overlaid.

**Figure 3:**
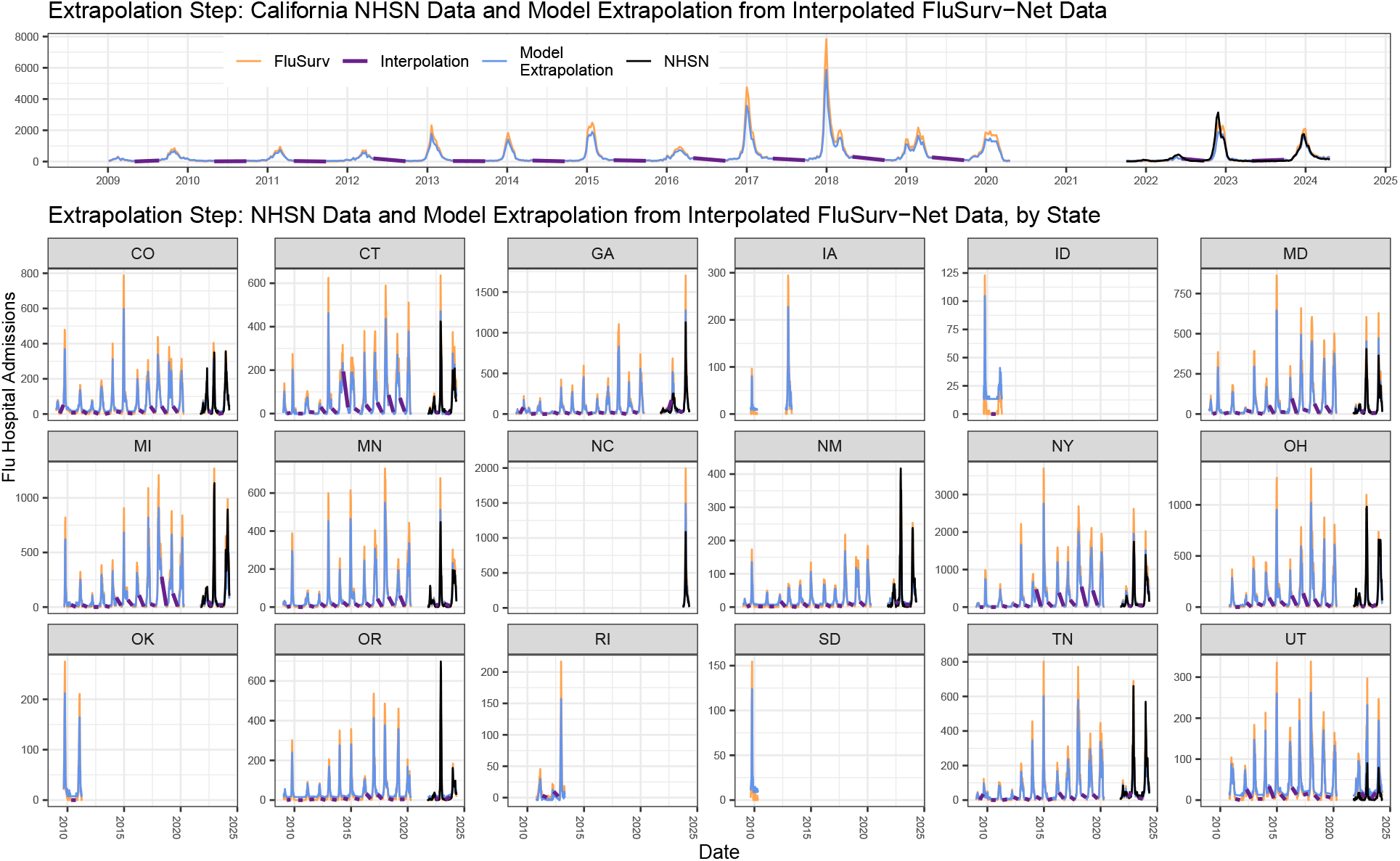
Extrapolation of FluSurv-Net data to NHSN data. The model extrapolation (blue line) is based on interpolated FSN data (purple line), state population size, and NHSN region. FSN records (orange) tend to be higher than NHSN reports (black), and the GLM extrapolation reflects this, often resulting in lower values than FSN records. Note that several states have no overlapping NHSN and FSN data, which is why variables like location were not used in the GLM despite their significance when both datasets were present.

#### 2.2.4 Imputation

After the extrapolation step, the dataset has a consolidated representation of flu hospitalizations comprised of the NHSN signal and the FSN data prepared using the steps described above. The dataset is still missing values for states that never or irregularly reported to FSN. With records dating back to 2009, the dataset contains 17,062 flu hospitalization records across 50 states, but there were an additional 22,888 missing hospitalization records. To complete the data augmentation and address the remaining missing gaps, we employed multiple imputation using the Multivariate Imputation by Chained Equations (MICE) algorithm (Van Buuren and Groothuis-Oudshoorn 2011). Prior to running the imputation, we added the ILINet data as an additional variable, which we queried using the cdcfluview R package (Rudis 2022) to collect state-level weekly counts of outpatient flu visits from 2010 to April of 2024. In addition to the hospitalization records, the dataset included incomplete cases for the ILINet outpatient visits (missing 11.39%) and the NHSN flu admission coverage rates (missing 76.97%). While these variables are rarely used in downstream analytics (e.g., forecasting models), they were significant predictors in the imputation model and were therefore simultaneously imputed by MICE. We also used the complete data fields, epiweek, epiyear, state population size, and HHS region, as predictors in the imputation. We did not explicitly include a state variable, because state population size perfectly covaries with state.

To select the best imputation methods for MICE, we evaluated eight plausible combinations by crossing two candidate methods for each of the three variables with missing data. For influenza hospitalizations, we tested Classification and Regression Trees (“cart”) and Predictive Mean Matching (“pmm”); for ILINet outpatient visits, we tested “pmm” and Normal Linear Regression Prediction (“norm.predict”); and for NHSN flu admission coverage, we tested “pmm” and Random Sampling from Observed Values (“sample”). These methods were selected based on the characteristics of each variable. Cart and pmm are well-suited for skewed or non-normally distributed count data like flu hospitalizations. Norm.predict assumes normality and was included as a benchmark for outpatient counts. Sample is appropriate for auxiliary variables with a high proportion of missing data or weak signal (Van Buuren 2018). To evaluate each combination, we randomly censored 20% of the flu hospitalization values and calculated the mean squared error (MSE) of the imputed values against the withheld “true” data. The best-performing combination used cart for flu hospitalizations and pmm for both ILINet and admission coverage. Cart imputes values by building decision trees that capture non-linear relationships and interactions, while pmm preserves the observed distribution by matching predicted values to actual cases in the data (Van Buuren 2018). Using this selected method set, we ran MICE with 150 imputations and 15 iterations to complete the dataset. Figure 4 illustrates the fully augmented dataset, which provides continuous estimates of influenza hospitalizations across all states from January 4, 2009 to April 21, 2024.

**Figure 4:**
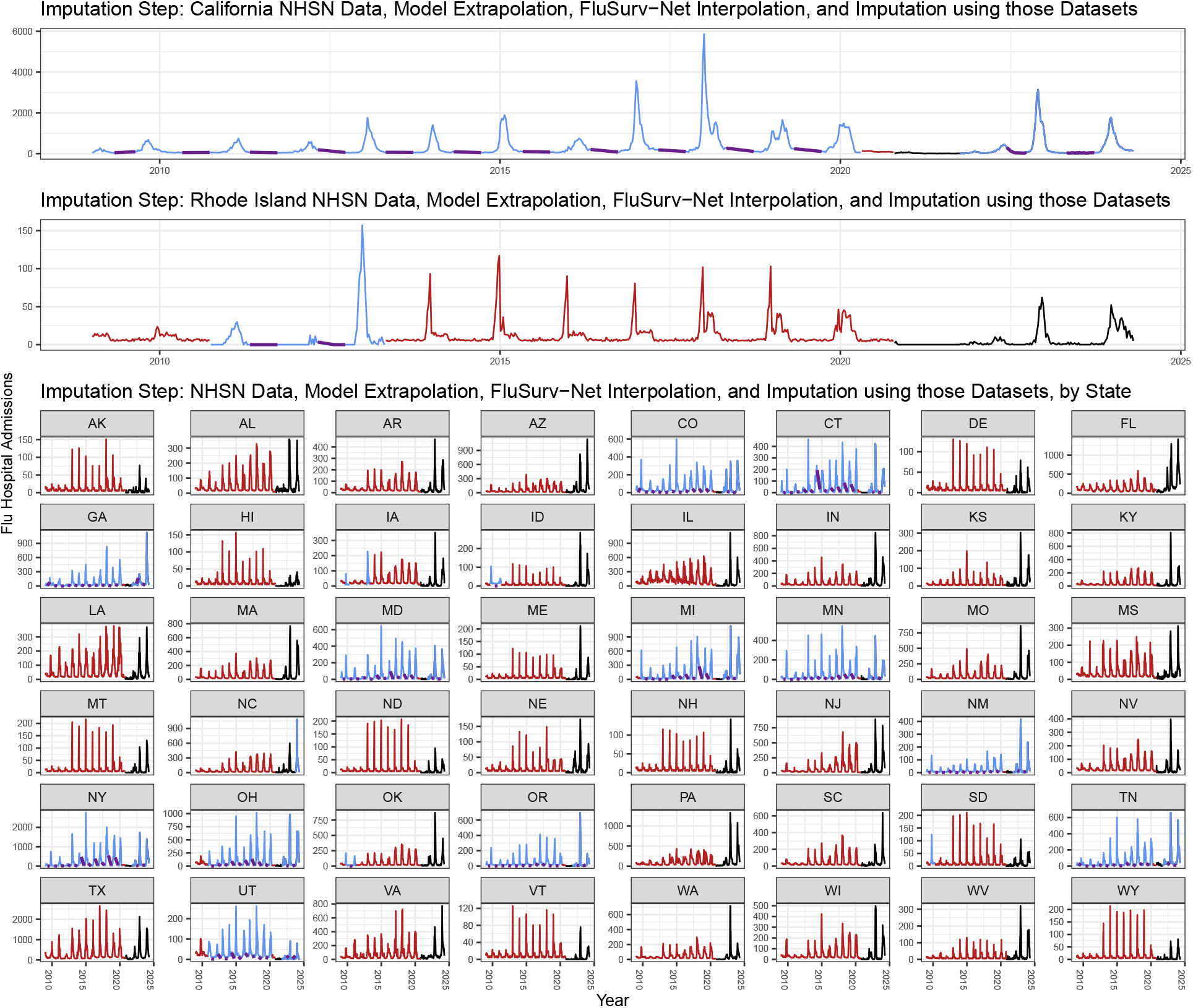
Imputation of remaining missing NHSN data. The final step in our augmentation completes the dataset, with imputed flu hospitalizations (red line), NHSN truth data (black line), extrapolated FSN data (blue line), and small interpolated gaps (purple line). Note that this figure is consistent in displaying California as an exemplar state (top panel) but also highlights Rhode Island. This comparison emphasizes the range in the amount of imputed data across states. California had a nearly complete flu hospitalization record between FSN data and NHSN data, with the exception of several months during 2020. On the other hand, Rhode Island’s record only included FSN data between 2011-2013 and NHSN data since 2021. Further, many states have never reported to FSN.

## 3 Performance and Method Variations

### 3.1 Imputation Accuracy

In extending the NHSN flu hospitalization data back in time, we have created a novel signal that hybridizes truly observed flu hospitalization data with informed estimates. For dates and locations for which there was no NHSN or FSN data previously available, we inherently lack truth data against which to compare the imputation results. This introduces challenges in assessing accuracy. In spite of these constraints, we attempted to demonstrate validity of our imputation procedure by censoring and then imputing a limited amount of more recent data for which we did have an NHSN signal. We performed this analysis by censoring data reported after October 2020 in two separate imputations: one in which data was censored in Tennessee and Utah, and the other in which it was censored in California and New Hampshire. When compared against the NHSN observed data, the seasonality of the imputations for the 2023-2024 flu season was fairly accurate, though less so for the seasons impacted by pandemic mitigation measures (e.g., mask mandates, social distancing, etc.). The accuracy of the imputed hospitalization burdens varied by state. Tennessee showed the most accurate burden, whereas the New Hampshire and Utah imputations slightly over-estimated the burden by less than 20 hospitalizations at seasonal peaks. Ultimately, without more extensive observed data, it is impossible to definitively assess the performance of the imputation. We emphasize that the imputed data were created to guide model training, hypothesis generation, and related activities. In terms of capturing a historical record of flu hospitalizations, the extensions to this time series do not have parity with true, reported signals.

### 3.2 Forecast Performance with Imputation

To demonstrate the utility of the augmented data set, we implemented a proof-of-principle, near-term forecast using a time-series approach. We based our forecast on an autoregressive integrated moving average (ARIMA) model. We implemented the ARIMA approach using the fiphde R package, which internally calls functions from the fable and fabletools packages (O’Hara-Wild, Hyndman, and Wang 2021a, 2021b). As part of this modeling procedure, the ARIMA performed a grid search of possible values for lag (p), differencing (d), and order (q) parameters, with ranges constrained 0:4, 0:4, and 0 respectively. The procedure was performed independently for each forecasting week and location to find the best fitting model, which was then used to generate forecasts for 4 weeks-ahead for the US and all 50 states for every week of the 2023-2024 flu season. We tested the model using variations of the dataset, each of which was iteratively masked to obscure training data for horizons that had not yet been eclipsed. The variations of training data included: the NHSN flu hospitalization dataset without any augmentation, the complete augmented dataset with hospitalizations dating back to 2009, a dataset truncated to exclude data before June 2010 (to account for the unusually severe flu season caused by the novel H1N1 virus (Centers for Disease Control and Prevention 2010)), a dataset excluding COVID-19 pandemic years, a dataset truncated to exclude data before June 2012 due to irregular reporting to FSN (i.e., several states reported inconsistently prior to this time, with large gaps or even multi-year periods without data), and several combinations of these exclusions. Note that for the dataset variation that excluded flu hospitalizations during the pandemic years, we removed data from the 2020-2021 and the 2021-2022 flu seasons, and shifted the data prior to May of 2020 forward by two years (i.e., the model assumes that the 2019-2020 flu season was the 2021-2022 flu season). We evaluated the performance of these variations using weighted interval scores (WIS), absolute error (AE), and the percent of forecasts that captured observed hospitalization counts within their 95% prediction intervals (95% coverage).

At the aggregated, national level, there were not substantial differences in performance between the forecasts using the original NHSN data (median WIS = 18.49, AE = 28, 95% coverage = 79.97%) and the variations of the imputed dataset (see Supplementary Figure S2). While the differences in performance were slight, the model trained on data that excluded flu hospitalizations prior to June 2010 demonstrated the best performance with a median WIS of 15.02, a median AE of 23.1, and capturing 83.16% of observed hospitalization counts. This suggests that excluding the 2009 flu season slightly enhances forecast accuracy. The dataset which excluded the COVID-19 pandemic and was truncated prior to June 2012 performed best in both the absolute error and the 95% prediction interval coverage metrics. All other configurations of the augmented dataset performed better than the original NHSN data across all three metrics, underscoring the benefits of the augmentation process in general (see Supplementary Figure S2).

For state-level forecasts, models that were trained on the full augmented dataset and those that were truncated prior to June of 2012 with pandemic years excluded generally performed best (see Supplementary Figures S3-S5). The model that used the full augmented dataset resulted in more state-level forecasts with the lowest median WIS values, closely followed by the model which used the 2012 truncated and pandemic excluded data. In most states, and across all three metrics, the original un-augmented dataset produced worse forecasts, with the exclusion of a few states with small population sizes. These findings emphasize the importance of balancing historical depth and the inclusion of significant events like the COVID-19 pandemic in training data to optimize the forecasting model’s performance across different states.

## 4 Discussion

The data augmentation approach we have developed and implemented provides a valuable resource for specifically improving flu hospitalization forecasting models and more generally demonstrating how an epidemiological time series may be extended. By combining data from FSN, NHSN, and ILINet, we have successfully created an estimate of state-level flu hospitalizations extending back to 2009. Flu hospitalization counts are particularly limited with only several years of reported data for all states. Furthermore, reporting mandates have shifted over time (U.S. Department of Health and Human Services 2023), and in the future, imputation procedures may be even more critical for this signal. An extended time series provides higher resolution, which also allows for forecasting methods that require more training data than was previously available (e.g., autoregressive neural network (Kandula and Shaman 2019; Turner, Hulme-Lowe, and Nagraj 2022)). Likewise, for time series methods that are currently being used (e.g., ARIMA approaches) we have demonstrated that access to this extended time series can improve forecast performance.

There are several limitations to the imputation approach and analysis described here. First, our decision to average the two New York jurisdictions from FSN could have introduced bias, as the FSN catchments for NY may capture hospitalizations in fundamentally different populations in the state. Furthermore, the choice of linear interpolation for addressing short gaps in the data might not be the most accurate method, but it was deemed sufficient given the available data and the context of the study. For the forecast evaluation, the inclusion of 2009 data, a year marked by a surge in flu cases due to the novel swine flu (Centers for Disease Control and Prevention 2010), might also skew the model training, though initial results indicate similar model performance with and without this data. Similarly, using data from 2020-2021 – years significantly affected by the COVID-19 pandemic – could impact model accuracy, although preliminary tests suggest that the model’s performance remains consistent regardless of the inclusion of 2020 data. Appropriate evaluation of forecast performance throughout the season is an area of active research, and the WIS, AE and coverage metrics we described are aggregated across all seasonal weeks. As such, the impact of extended time series on forecast performance at pivotal seasonal moments (e.g., initial rise, peak, etc.) may be obscured. Finally, we acknowledge that our demonstration analysis and the data augmentation procedure itself did not fully explore the impact of variability at each step. For example, while the extrapolation step is built on a GLM model, we only used the point estimate in downstream imputation, and we did not include ILINet data as a predictor in the model. Likewise, while we have standard error estimates for imputed hospitalization counts, we only used the point estimate as the input to our forecasting models in the performance analysis.

We anticipate that future work will focus on addressing the limitations above. We also recommend further validation of the imputation methods as the number of weeks of gold standard NHSN reporting grows over time. However, as noted above, changes to hospitalization reporting mandates may highlight an even more pressing need for data augmentation methods like that which we have presented here. We are aware of at least two other groups, both of which contribute to the CDC FluSight consortium, who have engaged in data fusion efforts to create augmented training data (Meyer et al. 2024; Ray et al. 2025). We have not yet evaluated those approaches in comparison to ours. Lastly, it is important to emphasize that the NHSN flu hospitalization dataset is by no means the only epidemiological signal that invites time series extension. We expect that our work may help provide a guide for other researchers to explore implementations of data augmentation approaches in other disease surveillance signals.

## 5 Conclusion

The procedure detailed here successfully combines FSN, NHSN, and ILINet datasets to address gaps and provide state-level estimates of flu hospitalizations dating back to 2009. The resulting dataset is anticipated to improve the performance of time series models used in forecasting and outbreak analytics, contributing to better preparedness and response strategies.

## Data Availability

All data and code produced are available online at: https://zenodo.org/records/16584391

https://zenodo.org/records/16584391

## 6 Acknowledgements

This work was supported in part by a subaward to Signature Science, LLC from the Council of State and Territorial Epidemiologists (CSTE) via the Centers for Disease Control and Prevention (CDC) Cooperative Agreement No. NU38OT000297.

The conclusions drawn and perspectives presented in this paper are solely those of the authors. However, the work would not have been possible without open and collaborative efforts from multiple entities. We acknowledge the following groups: the CDC for coordinating FluSight and providing guidance, interpretation, and dissemination of surveillance and forecast data; the CSTE for establishing collaborative networks through which forecasting groups can interact with one another and public health stakeholders; all participating teams in the FluSight network for their sustained contributions, innovative techniques, and commitment to openness through operational forecasting activities. We appreciate the willingness of members of the Reich Lab (UMass) and Machine Intelligence Group for the Betterment of Health and the Environment (Northeastern) to collaborate and share methods for augmenting influenza datasets.

## 7 Data Availability

The code and required data for the full augmentation process and the imputation results described here can be found at DOI 10.5281/zenodo.13146376.

