## Supplementary Figures for "An Imputation-Based Approach for Augmenting Sparse Epidemiological Signals"

### Supplementary Materials from: 'An Imputation-Based Approach for Augmenting Sparse Epidemiological Signals'

**Amy E. Benefield**
Signature Science, LLC


**Desiree Williams**
Signature Science, LLC

**VP Nagraj**
Signature Science, LLC

July 29, 2025

#### Abstract

Near-term disease forecasting and scenario projection efforts rely on the availability of data to train and evaluate model performance. In most cases, more extensive epidemiological time series data can lead to better modeling results and improved public health insights. Here we describe a procedure to augment an epidemiological time series. We used reported flu hospitalization data from FluSurv-NET, the National Healthcare Safety Network, and flu outpatient visits from ILINet to estimate a complete time series of flu hospitalization counts dating back to 2009. The augmentation process includes concatenation, interpolation, extrapolation, and imputation steps, each designed to address specific data gaps. We demonstrate the forecasting performance gain when the extended time series is used to train flu hospitalization models at the state and national level.

***K*eywords** Epidemiological modeling · Data augmentation · Time series · Flu hospitalizations · Imputation · Forecasting

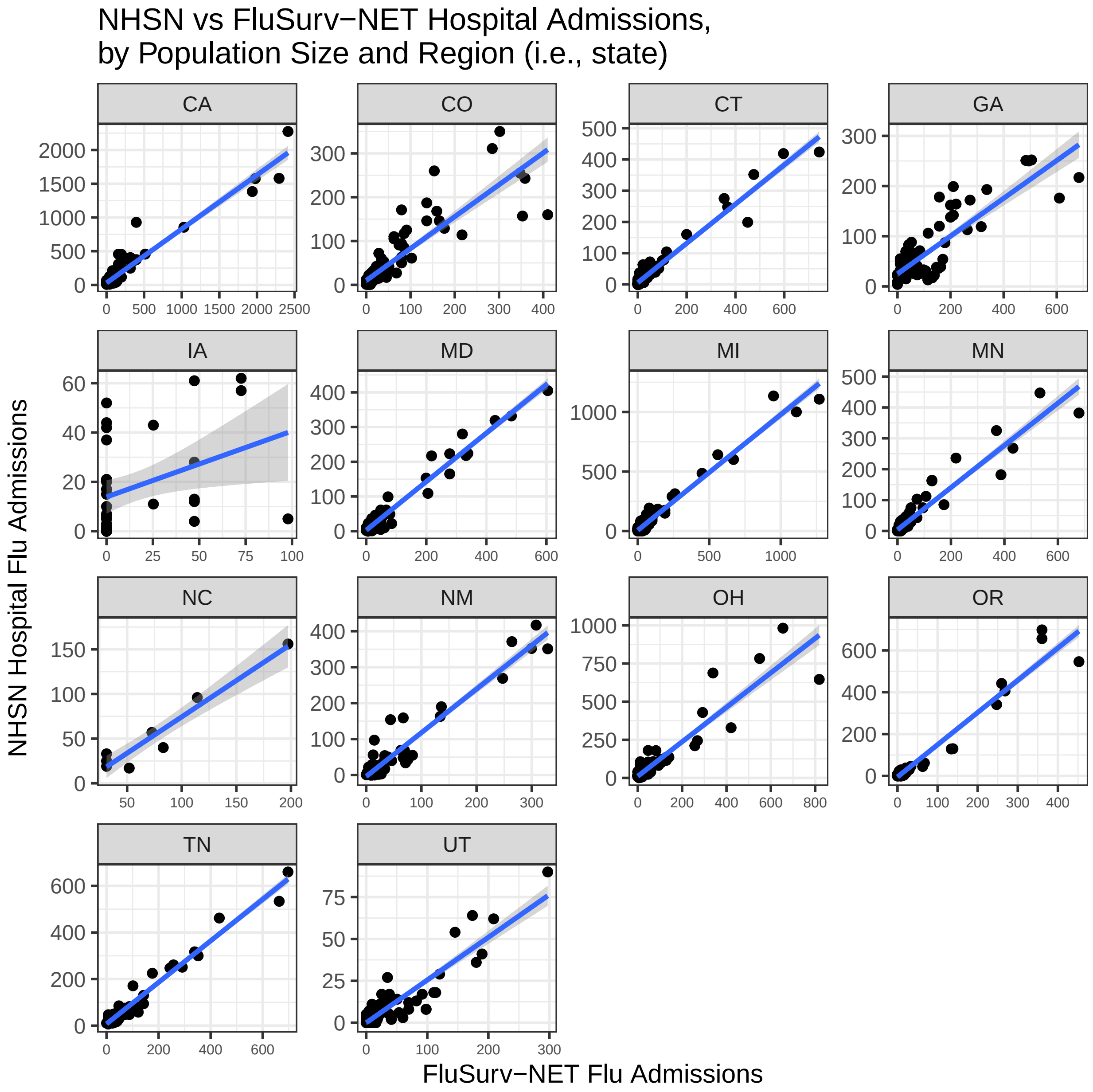


Figure 1: GLM model fit of NHSN flu hospitalizations versus FSN hospitalizations by population size and NHSN region. In this equation, state is functionally a composite variable that captures both NHSN region and population size, hence the figure is faceted by state. Both FSN hospitalizations and population size were significant predictors of NHSN hospitalizations ($p < 0.001$ for both), while NHSN region was marginally significant ($p = 0.128$). Model estimates, standard error, and point values are in blue, grey, and black, respectively. With the exception of Iowa, the model fit is fairly good. Iowa is likely a poorer fit due to its relatively smaller population size, fewer number of observations in FSN, and lack of additional representation from its HHS region (i.e., it was the only state in its HHS region reported in FSN).

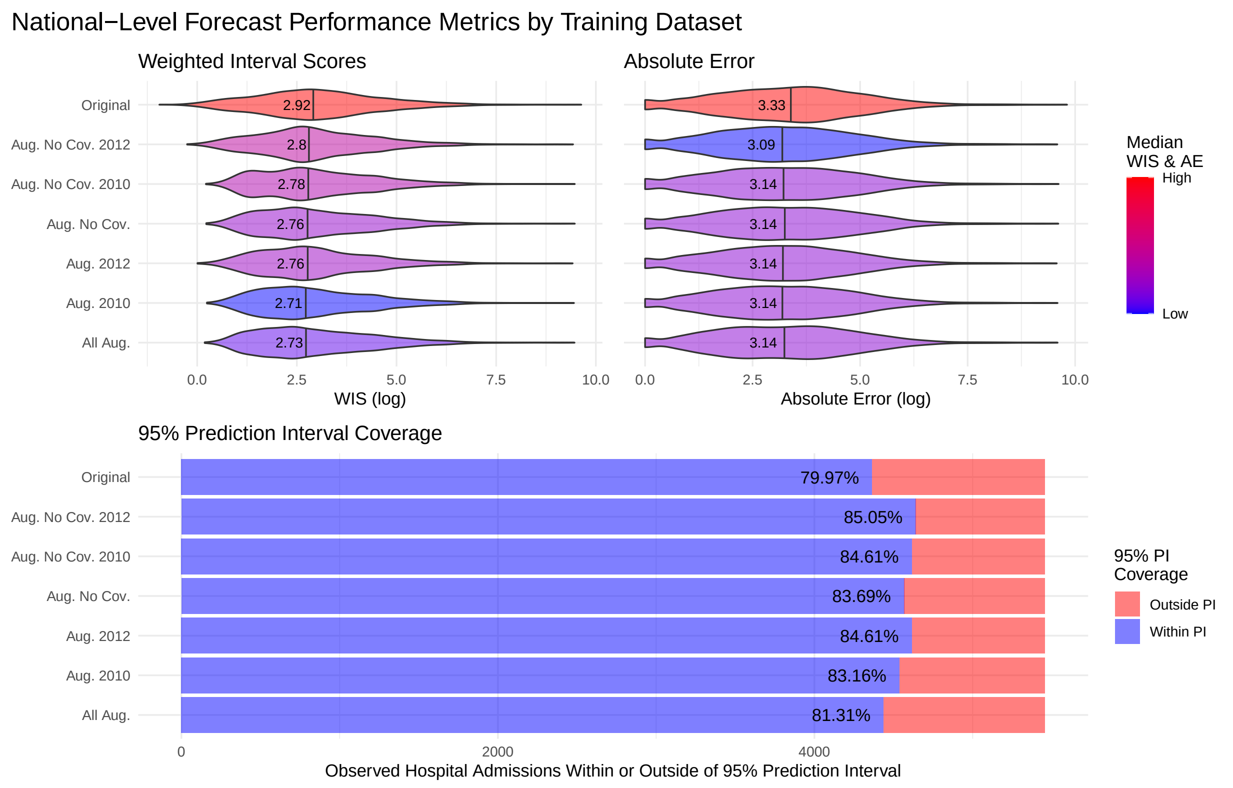


Figure 2: National-level forecast performance metrics by training dataset. The top left figure shows performance based on weighted interval scores (WIS), which is scaled such that a lower score indicates more accurate forecasts. There is not a great difference in performance across models, but the forecast generated using training data that was augmented and truncated before June 2010 performed the best and the original, NHSN-only training data performed the worst. The absolute error (AE) in the top right was similar in the performance variation, with the training dataset that was truncated before June of 2012 with the pandemic years excluded performing best. The bottom plot shows the percent of observed NHSN hospitalizations that were within the 95% prediction interval of the forecast model, with the best calibrated models being those that are closest to 95% in the coverage metric. [Key: Original = unaugmented NHSN data only; Aug. = augmented dataset; No Cov. = datasets where Covid-19 pandemic years were excluded; 2012 = augmented dataset truncated before June of 2012; 2010 = augmented dataset truncated before June of 2010; All Aug. = full augmented dataset with no exclusions or truncations]

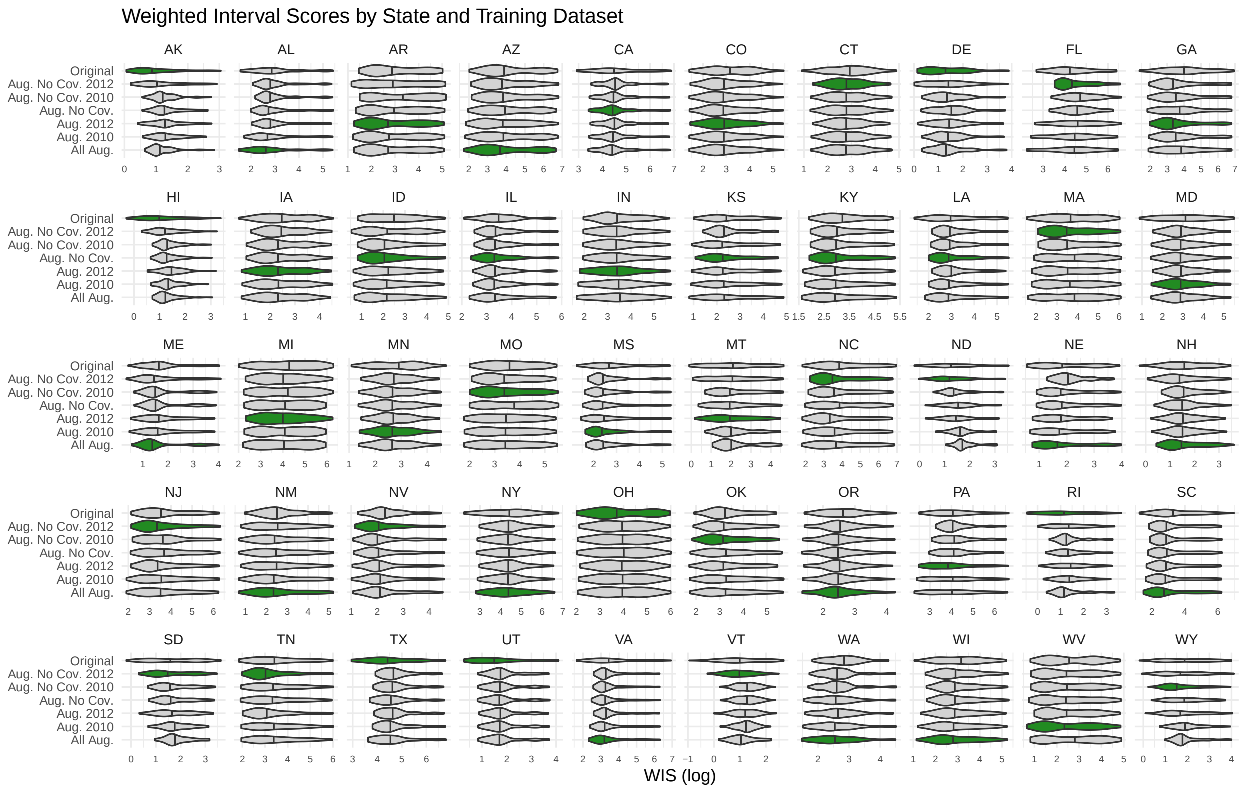


Figure 3: State-level weighted interval scores (WIS). The violins in green represent the datasets which produced the lowest median WIS (i.e., best performance). There is more variation in model performance based on training data at the state level (e.g., South Dakota). The dataset which performed the best in the most states (12 states) was the full augmented dataset. Conversely, the augmented dataset that was truncated before June of 2010, with pandemic years excluded, performed best in the fewest states (3 states). [Key: Original = unaugmented NHSN data only; Aug. = augmented dataset; No Cov. = datasets where Covid-19 pandemic years were excluded; 2012 = augmented dataset truncated before June of 2012; 2010 = augmented dataset truncated before June of 2010; All Aug. = full augmented dataset with no exclusions or truncations].

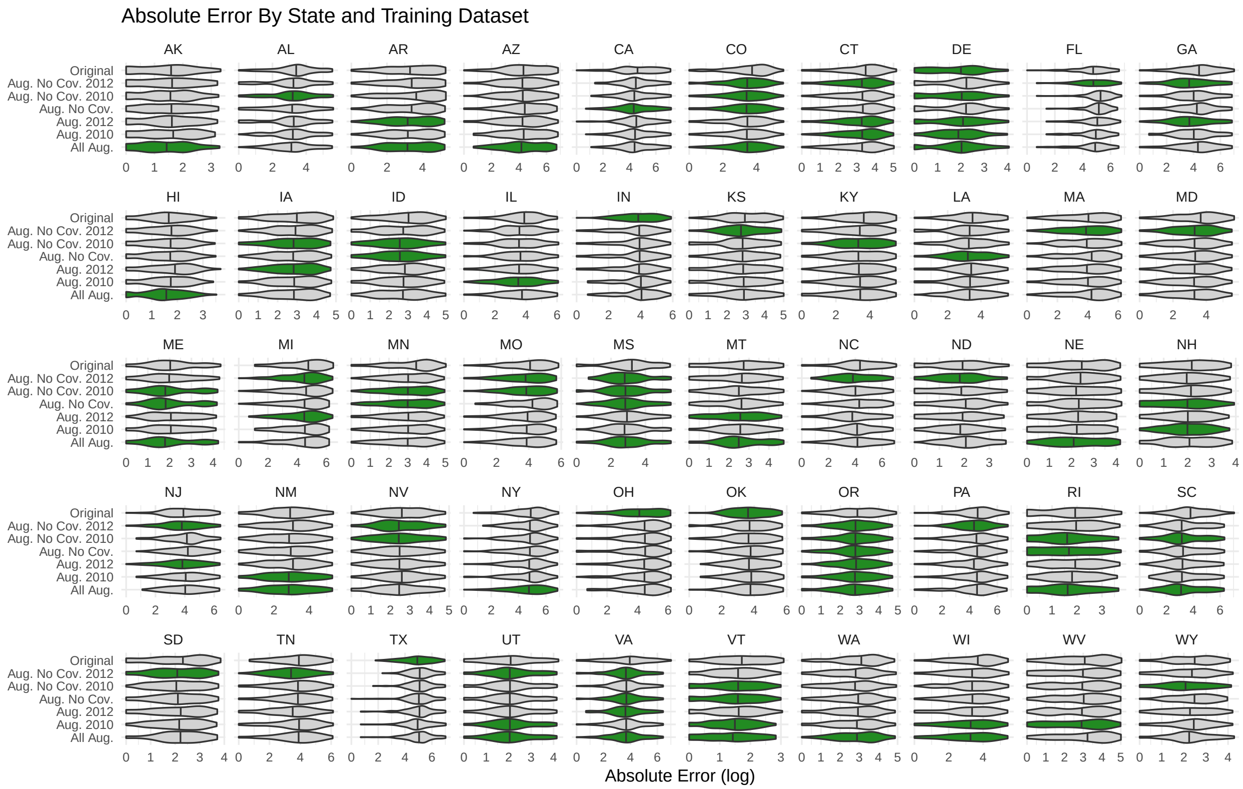


Figure 4: State-level absolute error (AE). The violins in green represent the datasets which produced the lowest median AE (i.e., best performance), and in some cases, there were ties (e.g., Colorado has a four-way tie for lowest median AE). The variation in model AE based on training data is about the same at the state and national levels (minimal). The datasets that performed the best in the most states (20 states each) were the full augmented dataset and the augmented dataset which was truncated before June of 2012 with the pandemic years excluded. Conversely, the original unaugmented NHSN-only dataset performed best in the fewest states (5 states). [Key: Original = unaugmented NHSN data only; Aug. = augmented dataset; No Cov. = datasets where Covid-19 pandemic years were excluded; 2012 = augmented dataset truncated before June of 2012; 2010 = augmented dataset truncated before June of 2010; All Aug. = full augmented dataset with no exclusions or truncations].

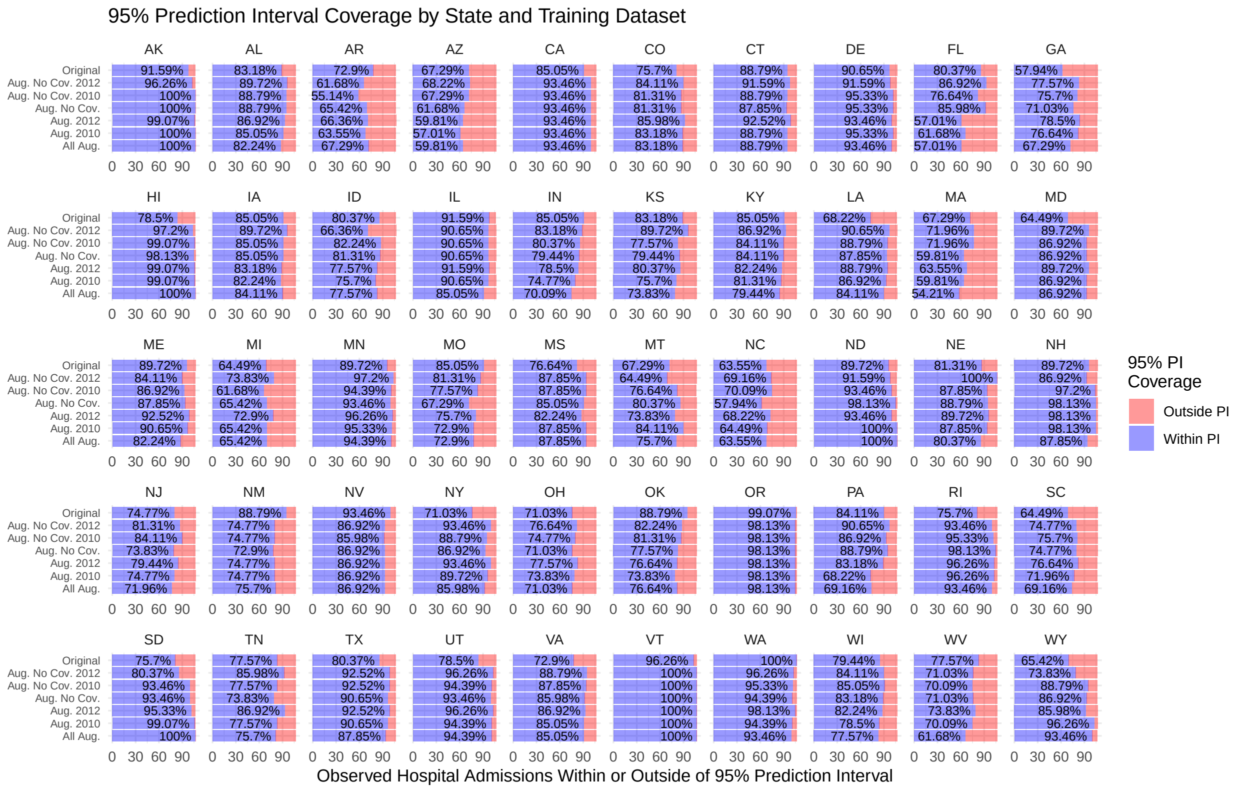


Figure 5: State-level percent of observed NHSN hospitalizations falling within the 95% prediction interval of the forecast by training dataset. Here the best calibrated models are those that are closest to 95% in the coverage metric. Calibration varied by model and state. However, in all but eight states, the inclusion of augmented data in some capacity improved calibration over the models that used original data. [Key: Original = unaugmented NHSN data only; Aug. = augmented dataset; No Cov. = datasets where Covid-19 pandemic years were excluded; 2012 = augmented dataset truncated before June of 2012; 2010 = augmented dataset truncated before June of 2010; All Aug. = full augmented dataset with no exclusions or truncations].
